# Women’s Body Mass Index trajectories from early pregnancy to one year postpartum and the rising burden of overweight and obesity over the last two decades in Bhaktapur, Nepal

**DOI:** 10.64898/2026.07.03.26357145

**Authors:** Manjeswori Ulak, Ram K. Chandyo, Adrian McCann, Ingrid Kvestad, Kjersti S. Bakken, Catherine Schwinger, Mari Hysing, Suman Ranjitkar, Merina Shrestha, Sudha Basnet, Tor A. Strand

**Affiliations:** Child Health Research Project, Department of Pediatrics, Institute of Medicine, Tribhuvan University, Kathmandu, Nepal; Siddhi Memorial Hospital, Bhaktapur, Nepal; Department of Community Medicine, Kathmandu Medical College, Kathmandu, Nepal; Bevital AS, Bergen, Norway; Regional Centre for Child and Youth Mental Health and Child Welfare, NORCE Norwegian Research Centre, Bergen, Norway; Women’s Clinic, Innlandet Hospital Trust, Lillehammer, Norway; Center for International Health, University of Bergen, Bergen, Norway; Department of Disease Burden, Norwegian Institute of Public Health; Department of Psychosocial Science, Faculty of Psychology, University of Bergen, Bergen, Norway; Department of Research, Innlandet Hospital Trust, Lillehammer, Norway

**Keywords:** BMI, Overweight, Obesity, Postpartum, Nepal

## Abstract

**Background and aims:** Maternal overweight and obesity are increasing worldwide, including Nepal. This study assessed BMI trajectories from early pregnancy to one year postpartum and trends in overweight and obesity over the past two decades in Bhaktapur, Nepal.

**Methods:** In the most recent study, BMI was measured in 800 Nepalese women at three time points: at early pregnancy, 6 and 12 months postpartum (2017 - 2021). The prevalence of undernutrition, overweight, and obesity was estimated using the World Health Organization and the Asian specific cut-offs. Long-term trends were assessed by comparing these findings with three population-based studies conducted in Bhaktapur between 2001 and 2021 among 2400 women at similar life stages.

**Findings:** Mean (SD) BMI increased from 23.7 (3.0) kg/m^2^ in early pregnancy to 26.1 (3.3) kg/m^2^ at 6 and 25.2 (3.3) kg/m^2^ and 12 months. The prevalence of overweight increased from 32.9% in early pregnancy to 48% at 6 months. Using the Asia-specific cut-offs, the prevalences were higher. Results from the three previous population-based studies demonstrated an upward trend where postpartum overweight increased from 11.4% in 2001–2002 to 44.6% in 2017 - 2021. The obesity prevalence rose from 1.8% to 10.9% during this period.

**Interpretation:** Overweight and obesity among Nepalese women have risen dramatically over the past two decades, with postpartum overweight increasing nearly fourfold and obesity more than sixfold. These findings highlight the need for interventions to prevent excessive weight retention and reduce adverse health outcomes.

## INTRODUCTION

Overweight and obesity (OWOB) are major public health challenges worldwide and are becoming increasingly prevalent in low- and middle-income countries (LMIC) (1). In marginalized settings, the double burden of malnutrition, where undernutrition coexists with overnutrition, is increasing due to nutritional transitions, including rapid changes in food quality and availability, and dietary habits (2).

The prevalence of obesity among adults aged 18 years and older has more than doubled between 1990 and 2022. The WHO further estimates that 43% of men and 44% of women worldwide are overweight (3). Prevalence varies by region, ranging from 31% in South-East Asia and Africa to 67% in the Americas. Overall, overweight and obesity have increased rapidly across South Asia, particularly among women and children, with women consistently exhibiting a higher burden of obesity than men (4). In Nepal, it has been reported that while the prevalence of undernutrition has declined over recent years, rates of OWOB have increased substantially (5). The 2019 Nepal STEPS Survey estimated that 25% of women aged 15-69 years were overweight and 5.3% were obese (6, 7). Similarly, the 2022 Nepal Demographic and Health Survey (NDHS) reported that among women aged 20–40 years, 26% were overweight and 8% were obese (8).

The postpartum period is a critical window for maternal health, as excessive weight retention after pregnancy is a major contributor to long-term OWOB and major risk factors for non-communicable diseases (NCDs), including diabetes mellitus, cardiovascular disease, hypertension, dyslipidemia, various cancers, and non-alcoholic fatty liver disease (9). NCDs currently account for approximately 74% of global deaths, according to the World Health Organization (WHO). From a Developmental Origins of Health and Disease (DOHaD) perspective, maternal metabolic health during and after pregnancy also influences the long-term health of offspring (10). Although OWOB has long been recognized as a major public health problem (11), the primary nutritional focus in many LMICs has, until recently, been on undernutrition. Consequently, longitudinal evidence on postpartum BMI trajectories and weight retention in South Asia remains limited. In the current article, we aimed to characterize women’s BMI trajectories from early pregnancy to 12 months postpartum using a longitudinal study of 800 pregnant women and to examine the changing burden of OWOB among women in Bhaktapur, Nepal over the past two decades, by comparing data across four community-based studies conducted in the same study setting.

## METHOD

### Study Setting

The study site was Bhaktapur municipality, located approximately 15 km east of the capital city, Kathmandu, at an altitude of 1,400 meters above sea level. According to the 2021 census, Bhaktapur municipality has a population of around 79,136, of whom about half (39,381) are women (12). The municipality is one of the most densely populated cities in Nepal. Most residents belong to the Newar ethnic group, although the municipality also includes migrant populations from other parts of Nepal, mainly Tamang (Lama) employed in carpet factories and other manual occupations. The main sources of income are agriculture, small-scale businesses, private or government service, and daily wage labor. The diet in the area is influenced by seasonality and local festivals. Most of the daily calories come from rice-based meals, and micronutrient intake has previously been shown to be generally inadequate (13).

### Study design

The most recent study is a prospective population-based cohort study nested within a double-blind, randomized, placebo-controlled trial (RCT) examining the effect of daily vitamin B12 supplementation (50 micrograms) during pregnancy and 6 months postpartum on child growth and neurodevelopment in early childhood (Clinical Trials registration number NCT03071666), conducted between 2017 and 2021 (14, 15). In this longitudinal study, women from the general population were recruited within 15 weeks of pregnancy between March 2017 and October 2020, and the one-year follow-up was completed in May 2022. A total of 800 pregnant women aged 20–40 years who planned to remain in the study area for at least the next 2 years were recruited. Women with high-risk pregnancy, acute or chronic diseases, anemia (hemoglobin < 7g/dL), or who were underweight (BMI <18.5) or obese (BMI ≥30) were excluded. Written informed consent, preferably in the presence of the husband, was obtained from all participants. The Nepal Health Research Council (NHRC) ethical review boards in Nepal (NHRC 253/2016) and the Regional Committee for Medical and Health Research Ethics (REC) in Norway (2016/1620/REK vest) approved the study.

To assess trends in OWOB over time, we compared findings from the current study with three previous community-based studies conducted in the same study setting. The first study (2000– 2001) examined micronutrient status among 500 non-pregnant women of childbearing age (13– 35 years) without acute or chronic disease (16). Ethical approvals were obtained from the Institute of Medicine, Tribhuvan University, Nepal (2064-10-17) and REC. The second study (2008–2009) was a cross-sectional study on the micronutrient status of 500 lactating women aged 15–45 years and their breastfed infants aged 2–11 months recruited through two-stage cluster sampling (17). Ethical approval was obtained from the Institute of Medicine, Tribhuvan University, Nepal and REC. The third study (2015–2017) was a community-based, double-blind RCT of daily vitamin B12 supplementation among 600 mildly stunted infants aged 6–11 months (18). At the time of enrolment, we also measure the height and weight of the participants’ mothers using a standard protocol to examine maternal nutritional status. Ethical approval was obtained from the NHRC (NHRC; #233/2014) and REC (REC; #2014/1528).

In all four studies, except the first study (2001-2002), the women were in the postpartum period between 2 months and 12 months, and measurements were taken using a standard method by trained field workers.

### Procedures

For the current study and three previous studies, participating women were interviewed to collect socio-demographic and household information, including education, occupation, and income, as well as clinical and antenatal information at enrollment.

#### Anthropometric measurement

Trained field workers measured weight and height using standardized methods in all studies. Weight was measured using electronic scales (Salter/HoMedics Group, UK, and Seca, Germany) with a precision of 100 g, while height was measured using a stadiometer (Prestige, Hardik Medi Tech, India) with a precision of 1 mm. Participants were asked to remove shoes and heavy clothing before the anthropometric measurements. The weighing scales were regularly calibrated using certified standard weights (1 kg, 5 kg, and 10 kg), and the stadiometer was calibrated by verifying the measurements against a certified standard length using a measuring tape.

### Outcome

BMI was calculated as weight in kilograms divided by height in meters squared (kg/m^2^) (19). We categorized BMI according to the WHO recommendation for underweight (BMI <18.5 kg/m^2^), normal weight (BMI 18.5–24.9 kg/m^2^), overweight (BMI 25–29.9 kg/m^2^), and obesity (BMI ≥30 kg/m^2^) (11). As South Asian populations have a higher risk of metabolic diseases at lower BMI levels compared with Western populations, we also used WHO-recommended Asian-specific cut-offs: underweight (BMI <18.5 kg/m^2^), normal weight (18.5–22.9 kg/m^2^), overweight (BMI 23–24.9 kg/m^2^), and obesity (BMI ≥25) (20).

### Statistical analysis

Descriptive statistics were used to summarize participants’ sociodemographic characteristics. Continuous variables were reported as means and standard deviations (SDs), and categorical variables as numbers (n) and percentages (%). The BMI of women is reported as mean (SDs). Transitions between BMI categories from early pregnancy to 6 and 12 months postpartum were visualized using Sankey diagrams (Stata package Sankey-plot). The diagram illustrates the number of participants moving between BMI categories (i.e., normal weight, underweight, overweight, and obese) over time. The WAMI index was used as a measure of socioeconomic status and calculated based on four components: access to water and sanitation, household assets, maternal education, and household income. WAMI scores range from 0-1 with higher scores indicating higher socioeconomic status.

In the longitudinal sample of 800 pregnant women, logistic regression analyses were performed to examine factors associated with overweight and obesity at 12 months postpartum. We present unadjusted and adjusted odds ratios (ORs) with 95% confidence intervals (CIs). We derived the final adjusted model using purposeful selection of variables according to the procedures suggested by Hosmer and Lemeshow (21). Adjustment variables considered relevant were based on previous literature or variables associated with the outcome at a P-value <0.20.

Maternal BMI distributions were compared across study periods using covariate-standardised kernel density plots. Individual-level data from the included studies were harmonised and stacked into a single dataset. Maternal BMI was used as the outcome variable, and the study period was coded as a categorical variable. To account for differences in the distribution of maternal age and time since last childbirth across study periods, we used inverse probability weighting. All statistical analyses were performed using Stata (version 16; StataCorp LLC, College Station, TX, USA).

## RESULTS

### Participant characteristics

**Table 1** presents the demographic and socioeconomic characteristics of women from the most recent longitudinal study. The participants had a mean age of 27.5 ± 4.0 years, with 45.5 % aged 25–29 years. Most women had secondary education (70%), while 23% had attained higher education and 7% had little or no schooling. The majority belonged to the Newar ethnic group (78.3%), most women (65.3%) lived in joint families, one-fourth (24.8%) in rented houses, and 26.8% had shared kitchen–bedroom spaces. Few families owned land (5.5%) or received remittances from abroad (10.6%). The mean WAMI score was 0.66 ± 0.13. At enrolment, almost half (48.7%) of women were first-time pregnant (primiparous), and 9% of all births were preterm. Caesarean delivery was reported in almost half (47%) of the participants, and a mean birth weight of 3015 ± 457 g. Exclusive breastfeeding was on average shorter than recommended, with 29.6% practicing it for less than 3 months, and only 13.8% for six months or more.

**Table 1.** Demographic and Household Characteristics in a longitudinal cohort of 800 Women During Early Pregnancy and the Postpartum Period in Bhaktapur, Nepal.

| Demographic and socioeconomic features | N (%) |
| --- | --- |
| Women's age in years, mean ± SD | 27.5 ± 4.0 |
| **Age, year** |  |
| 20–24 | 195 (24.4%) |
| 25–29 | 365 (45.5%) |
| 30–35 | 198 (24.7%) |
| >35 | 43 (5.4%) |
| **Education:** |  |
| Illiterate or Primary (1–5 grade) | 55 (6.9%) |
| Secondary (6–12 grade) | 560 (70.0%) |
| Higher (>12 grade) | 185 (23.1%) |
| **Occupation of women:** |  |
| Homemaker/Agriculture | 283(35.4%) |
| Service (monthly salary) | 279 (34.9%) |
| Self-small business or daily wage worker | 238(29.7%) |
| **Caste/ethnicity:** |  |
| Newer | 626 (78.3%) |
| Brahmin/Chhetri | 77 (9.6%) |
| Indigenous group /others | 97 (12.1%) |
| **Socio-economic features:** |  |
| Live in joint family | 522 (65.3%) |
| Live in rented house | 198 (24.8%) |
| Kitchen and bedroom in the same room | 216 (26.8%) |
| Family having own land | 44 (5.5%) |
| Receiving remittance from abroad | 85 (10.6%) |
| Wealth index (WAMI), mean ± SD | 0.66 ± 0.13 |
| **Perinatal information at enrollment** |  |
| Parity, Primiparous | 390 (48.7%) |
| Secundiparous | 376 (47.0%) |
| Multiparous | 34 (4.3%) |
| Caesarian section delivery | 361 (47.1%) |
| Birth weight of child, grams (mean ± SD) | 3015 ± 457 |
| Child born at gestational age (among 767) |  |
| Preterm (<37 wks) | 70 (9.1%) |
| Full term (37–40 wks) | 653 (85.1%) |
| Post term (>40 wks) | 44 (5.8%) |
| Duration of exclusive breastfeeding: |  |
| <3 months | 219 (29.6%) |
| 3–5 months | 217 (29.4%) |
| ≥6 months | 102 (13.8%) |

The characteristics of participants in the three previous studies are presented in **Supplementary Table 1**. Comparing the participants in the most recent study, the educational attainment is improved, with the proportion of women completing grade 10 or higher increasing from 20% in 2001–2002 to 72.9% in 2017–2021. The proportion engaged in homemaking or agriculture has declined from 41% to 35% in the same period. Furthermore, household characteristics have changed, where the proportion of women living in joint families increased from 51.6% to 65.3%, while average family size decreased from 6.5 ± 3.1 to 5 ± 2.5 members (**Supplementary Table 1**).

### Maternal BMI throughout early pregnancy to postpartum in the most recent longitudinal study

Mean BMI increased from 23.7 ± SD: 3.0 in early pregnancy to 26.1 ± 3.3 at 6 months postpartum, slightly declining to 25.2 ± 3.3 at 12 months. A marked shift in BMI categories was observed, and the prevalence of normal weight and OWOB varied substantially depending on the BMI classification used (Table 2 and supplementary Table 2). Based on WHO BMI classifications, 67% of women had a normal BMI at baseline. The proportion with normal BMI decreased to 38.3% at 6 months postpartum, while the prevalence of overweight and obesity increased to 48.0% and 13.2%, respectively. At 12 months postpartum, the prevalence of overweight and obesity was 41.2% and 8.6%, respectively **(Table 2)**.

**Table 2.**
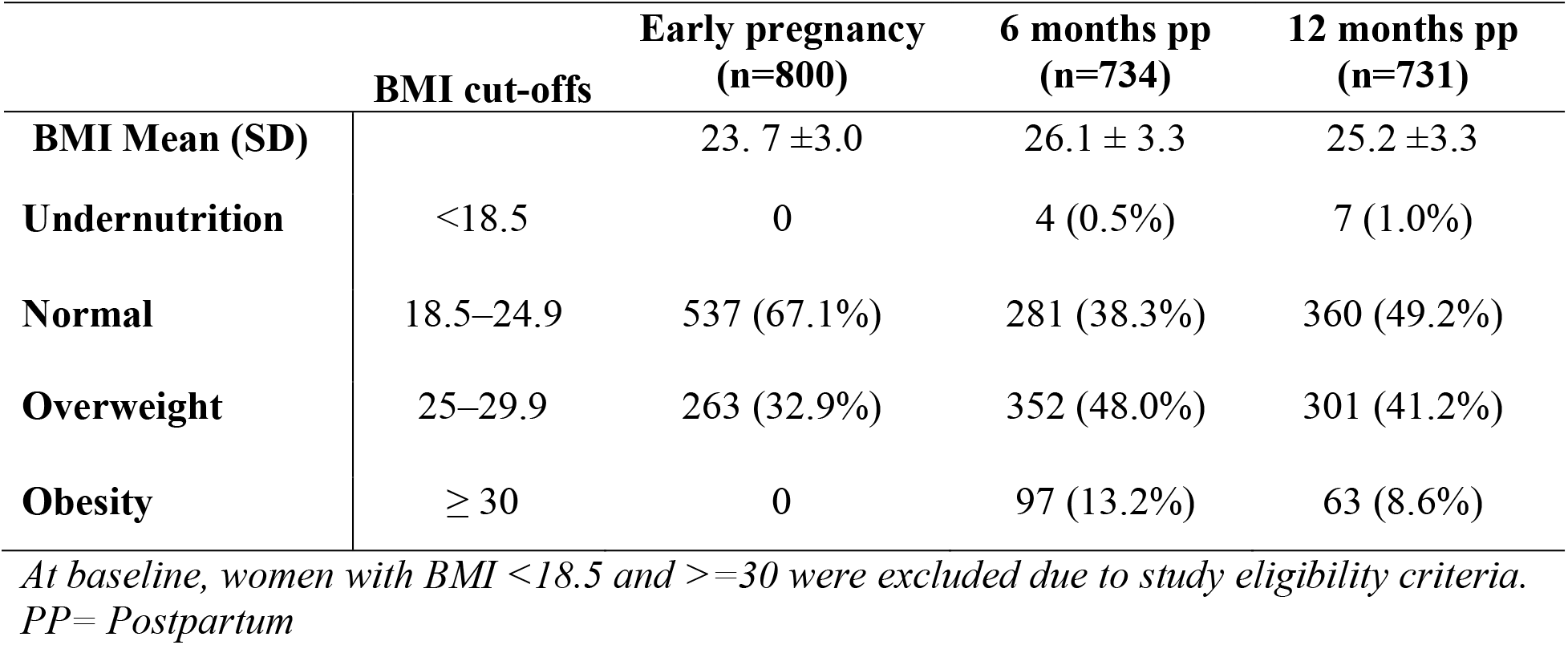
Prevalence of overweight and obesity in pregnant in a longitudinal cohort of 800 Women During Early Pregnancy and the Postpartum Period in Bhaktapur, Nepal.

Applying South Asian BMI cut-offs resulted in markedly higher estimates of adiposity. In the first trimester, 32.8% of the women were classified as obese (BMI ≥25). This proportion increased sharply to 61.2% at 6 months postpartum and remained high at 49.8% at 12 months postpartum **(Supplementary table 2)**.

### Transitions in women’s BMI categories from early pregnancy to 12 months postpartum

The Sankey diagram illustrates the tracking of BMI categories over time (**Figure 1**). Most women were in the normal weight and overweight categories during the first trimester of pregnancy. Notable transitions toward higher BMI categories, particularly from normal to overweight and from overweight to obesity, were observed at 6 months postpartum. By 12 months postpartum, most women remained in the overweight category, with some transitioning to or remaining in the obese category, suggesting an increased burden of overweight and obesity in the cohort.

**Figure 1.**
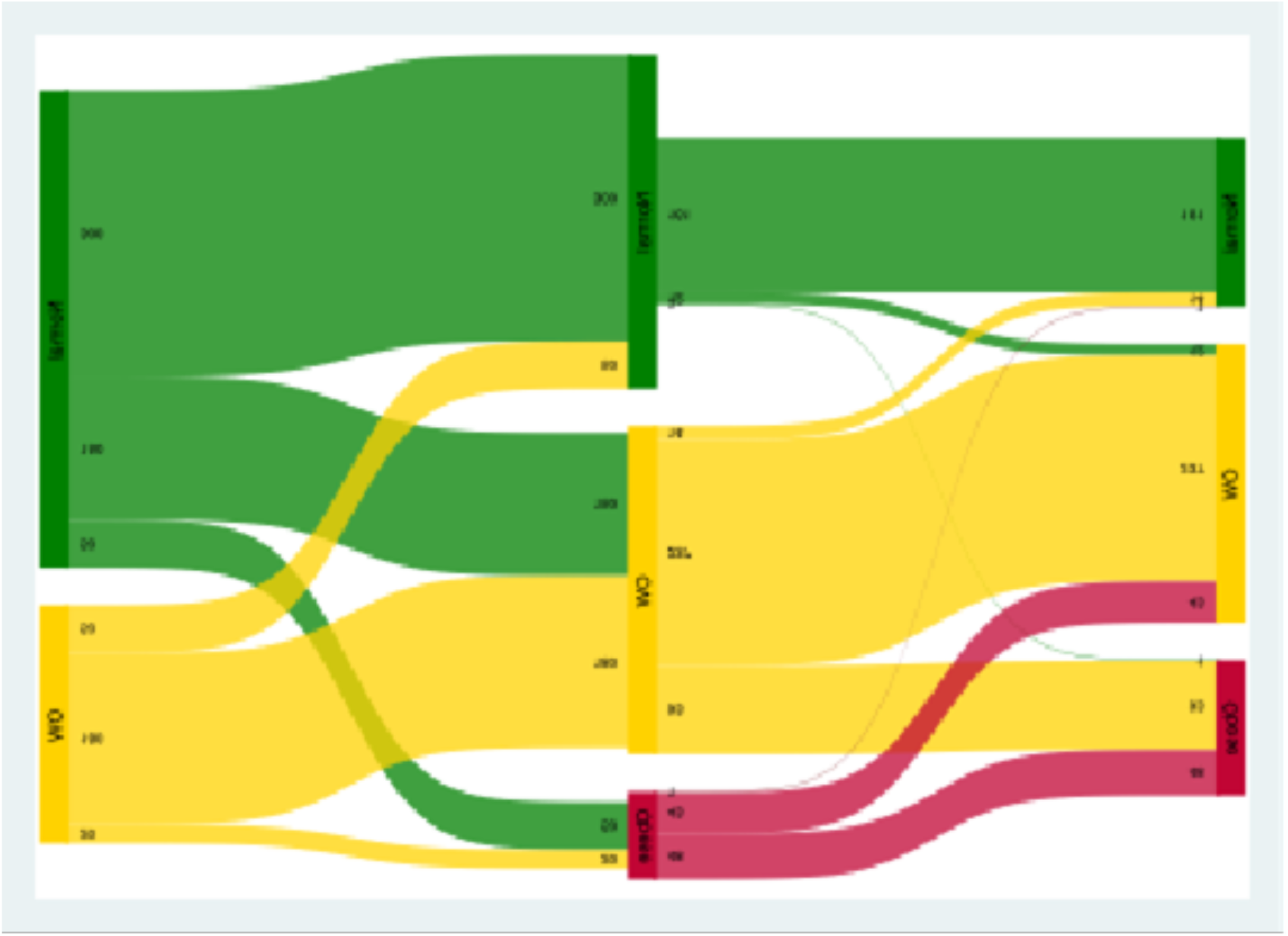
Overweight and Obesity of women at baseline, 6, and 12 months postpartum according to the cut-off BMI by the WHO definition (Sankey graph)

**Table 3** presents the crude ORs and adjusted ORs (AORs) for factors associated with overweight or obesity among the women at 12 months postpartum. Increasing age was associated with higher odds of overweight or obesity. Compared with women aged 20–24 years, women aged 25–29 years (AOR: 1.5; 95% CI: 1.1–2.2), 30–35 years (AOR: 2.1; 95% CI: 1.3–3.4), and >35 years (AOR: 4.1; 95% CI: 1.8–9.3) had progressively greater odds of overweight or obesity. Women in the highest wealth quartile (WAMI) also had significantly higher odds of overweight or obesity compared with those in the lowest quartile (AOR: 1.9; 95% CI: 1.1–3.3). While multiparous women had higher crude odds of overweight or obesity (OR: 1.8, P <0.010), the association was not statistically significant after adjustment for potential confounders (AOR: 1.4, P value 0.59). Education level and family type were not significantly associated with overweight or obesity in the adjusted model.

**Table 3.** Factors associated with overweight or obesity among women 12 months postpartum in a longitudinal study in Bhaktapur, Nepal.

|  | Crude |  |  | Adjusted* |  |  |
| --- | --- | --- | --- | --- | --- | --- |
|  | OR | 95% CI | P value | OR | 95% CI | P value |
| **Age, year** |  |  |  |  |  |  |
| ▪ 20–24 | Ref. |  |  | Ref. |  |  |
| ▪ 25–29 | 1.6 | 1.1–2.3 | 0.01 | 1.5 | 1.1–2.2 | 0.04 |
| ▪ 30–35 | 2.6 | 1.7–3.9 | <0.01 | 2.1 | 1.3–3.4 | 0.00 |
| ▪ >35 | 5.1 | 2.4–11.2 | <0.01 | 4.1 | 1.8–9.3 | 0.00 |
| **Education** |  |  |  |  |  |  |
| ▪ Secondary (6–12 grade) | Ref. |  |  | Ref. |  |  |
| ▪ Illiterate or Primary (1–5 grade) | 1.7 | 1.0–3.2 | 0.06 | 1.7 | 0.8–3.3 | 0.21 |
| ▪ Higher (>12 grade) | 1.1 | 0.8–1.6 | 0.49 | 0.9 | 0.6–1.4 | 0.60 |
| **Family type** |  |  |  |  |  |  |
| ▪ Nuclear | Ref. |  |  | Ref. |  |  |
| ▪ Joint | 0.8 | 0.6–1.1 | 0.16 | 0.9 | 0.6–1.3 | 0.66 |
| **Wealth index (WAMI)** |  |  |  |  |  |  |
| ▪ Q1 (Lowest) | Ref. |  |  | Ref. |  |  |
| ▪ Q2 (Middle) | 1.2 | 0.2–1.8 | 0.42 | 1.5 | 0.9–2.3 | 0.09 |
| ▪ Q3(Middle) | 1.0 | 0.7–1.6 | 0.82 | 1.5 | 0.9–2.4 | 0.12 |
| ▪ Q4 (Highest) | 1.4 | 0.91–2.1 | 0.12 | 1.9 | 1.1–3.3 | 0.01 |
| **Parity:** |  |  |  |  |  |  |
| ▪ Primipara | Ref. |  |  | Ref. |  |  |
| ▪ Multiparous | 1.8 | 1.3–2.4 | <0.01 | 1.4 | 0.9–1.9 | 0.59 |

\*Adjusted model includes all predictors

### Secular Trends in BMI or Nutritional Status Among Women in Bhaktapur, Nepal, over the last 20 years (2001–2021)

**Table 4** shows a clear increasing trend in BMI among women of childbearing age and postpartum over the past two decades using WHO BMI cut-offs. The mean BMI has steadily increased from 21.8 ±3.0 in 2001–2002 to 22.5 ± 3.1 in 2007–2008, further rising to 23.7 ±3.6 in 2015–2017, and reaching 25.6 (±3.3) in 2017–2021. This indicates a progressive increase in body weight over time. The prevalence of overweight (25–29.9 kg/m^2^) has increased consistently, rising from 11.4% in 2001–2002 to 44.6% in 2017–2021. Similarly, obesity (≥30 kg/m^2^) has shown a notable upward trend, increasing from 1.8% to 10.9% over the same period. Notably, these increases occurred even though women with obesity at enrolment were excluded from the present study.

**Table 4.** Trends in BMI according to WHO cut-off among women of childbearing age or during lactation in four studies conducted the last 20 years in Bhaktapur, Nepal.

| Year | Total Sample | Period | BMI Mean (SD) | BMI (%) |  |  |  |
| --- | --- | --- | --- | --- | --- | --- | --- |
|  |  |  |  | <18.5 | 18.5–24.9 | 25–29.9 | ≥30 |
| 2018–2021 | 731 | Postpartum 6–12 months | 25.6 (±3.3) | 5 (0.7%) | 320 (43.8%) | 326 (44.6%) | 80 (10.9%) |
| 2015–2017<br>Third study | 600 | Postpartum 6–11 months | 23.7 (±3.6) | 34 (5.7%) | 357 (59.5%) | 175 (29.2%) | 34 (5.7%) |
| 2007–2008<br>Second study | 500 | Postpartum 2–11 months | 22.5 (±3.1) | 23 (4.6%) | 392 (78.4%) | 70 (14%) | 15 (3%) |
| 2001–2002<br>First study | 500 | Childbearing age | 21.8 (±3) | 55 (11%) | 379 (75.8%) | 57 (11.4%) | 9 (1.8%) |

Using South Asian BMI cut-offs (**Supplementary Table 3**), the prevalence of obesity (BMI ≥25 kg/m^2^) increased markedly from 13.2% in 2001–2002 among women of childbearing age to 61.2% among postpartum women in 2017–2021. BMI distributions by study are shown in Supplementary Figures 1 (covariate-adjusted) and 2 (unadjusted). Covariate adjustment was not possible for the 2001 survey because relevant covariate data were unavailable.

## DISCUSSION

In a longitudinal cohort from peri-urban Bhaktapur, Nepal, BMI rose from early pregnancy through the postpartum period, peaking at 6 months and remained elevated at 12 months. Nearly half of the women were classified as overweight and >10% as obese in early postpartum. Using data across study periods over the last 20 years, show an almost fourfold increase in overweight and more than sixfold increase in obesity among women of childbearing age and in the postpartum period from the same study setting.

### Weight trajectories among women in childbearing age and postpartum

Our findings that postpartum weight retention remained elevated through 12 months postpartum are consistent with prospective studies from South Asia, particularly India, which have reported persistent postpartum weight retention during the first postpartum year (22), which identify the postpartum months as the period of greatest net weight retention. Twelve months postpartum is commonly used in epidemiological studies as a key time point for evaluating postpartum weight retention and maternal weight recovery (23). The persistence of elevated BMI at 12 months postpartum in our study also aligns with global evidence showing that many women fail to return to their pre-pregnancy weight within the first year after childbirth (24). The result from the longitudinal cohort is underlined by findings across studies over the two decades, showing a steady increase in the prevalence of OWOB in women of childbearing age and the postpartum period. These increasing trends are of clinical and public health concern, as postpartum weight retention can lead to long-term obesity and cardiometabolic disease, and maternal obesity increases the risk of adverse health outcomes in both mothers and children. These findings support the need for interventions targeting the early postnatal period to prevent long-term weight gain.

### Predictors of postpartum weight

In the longitudinal cohort, we found that older maternal age and higher household wealth were independently associated with overweight/obesity at 12 months postpartum. These reversed socioeconomic gradients have been reported in Nepal and neighboring countries previously, where increased wealth correlates with higher OWOB prevalence among women (25) (11), potentially signaling greater access to energy-dense foods and more sedentary occupations. Age effects likely reflect cumulative weight retention and age-related metabolic changes and are consistent with studies from South Asia and Nepal (1, 11, 26).

The postpartum rise in BMI plausibly reflects multiple interacting drivers: biological postpartum weight retention, reduced physical activity, and culturally prescribed postpartum diets and rest practices that encourage energy-dense intake and limited mobility (31). Qualitative and observational studies from South Asia document these traditions and their likely impact on postpartum energy balance and behavior (24). Similarly, studies from India highlight that social and cultural beliefs strongly influence postpartum diet and physical activity behaviors, with many women reporting reliance on family advice and limited awareness of evidence-based weight management strategies (27).

### Secular change and public-health implications

Placed alongside earlier local studies and national surveys, our data are consistent with a broader nutritional transition in Nepal: undernutrition has declined while overweight and obesity have increased among women of reproductive age, transforming the population risk profile. The most recent Nepal Demographic and Health Survey also confirms this transition, reporting that OWOB now affects a substantial proportion of women of reproductive age, surpassing undernutrition in many settings (8). National data from Nepal also show a striking rise in prevalence: overweight among women increased from 1.8% in 1996 to 19.7% in 2016, with obesity alone rising from 0.2% to 4.1% (28). Our findings extend this evidence by demonstrating that excess weight is particularly concentrated during the postpartum period, suggesting that reproductive transitions may amplify population-level trends. This pattern mirrors broader South Asian evidence, where rapid urbanisation, dietary shifts, and reduced physical activity have driven a sharp increase in obesity among women (29). Despite this, maternal health services in South Asia remain largely focused on pregnancy, with limited structured postpartum follow-up to support weight management (30). As postpartum weight retention can seed long-term maternal obesity and cardiometabolic risk, early recognition and intervention, including promotion of balanced diets and physical activity, are critical to prevent long-term health consequences for both mothers and offspring.

### Strengths and limitations

Strengths include the longitudinal design and ability to compare individual trajectories with regional secular data; major limitations are the exclusion at baseline of women with BMI <18.5 or ≥30 kg/m2 (which constrains generalizability) and the lack of measured diet and physical activity, so residual confounding by these behaviors cannot be excluded. We have therefore been cautious when inferring behavioral mechanisms. The sampling procedures differed across the studies included in this analysis; therefore, the observed 20-year trend in maternal BMI should be interpreted with some caution. The studies were not designed as repeated cross-sectional surveys with identical sampling frames, and variations in recruitment procedures, eligibility criteria, assessment timing, and participant characteristics may have contributed to differences in the observed BMI distributions. Thus, part of the apparent secular change could reflect differences in study design rather than true population-level change.

To assess whether differences in key maternal characteristics explained the observed patterns, we generated covariate-standardized BMI density plots using inverse probability weighting. These plots were standardized for maternal age and time since last pregnancy/childbirth, as these variables differed between studies and are plausibly related to maternal BMI. Adjustment for these covariates did not substantially alter the BMI distributions, as shown in Supplementary Figures 1 and 2. This finding suggests that the overall pattern of increasing BMI across study periods was not primarily driven by differences in maternal age or time since pregnancy.

## Conclusion

This study demonstrates a clear and sustained increase in BMI during the postpartum period, embedded within a two-decade-long secular rise in OWOB in Bhaktapur, Nepal. The findings highlight postpartum women as a high-risk group within an ongoing nutritional transition and highlight the need for integrated maternal nutrition strategies that extend beyond pregnancy to address the growing burden of obesity.

## Data Availability

The datasets generated and analyzed during the current study are not publicly available because they contain sensitive participant information and are subject to ethical and institutional restrictions. De-identified data may be made available upon reasonable request subject to approved by the Nepal Health research Council (NHRC) and Regional Committee for Medical and Health Research Ethics in Norway.

## Contributors

MU: conceptualization, study design, data collection, supervision, data analysis, and wrote the original draft of the manuscript. TS: the study design, data analysis, interpretation of the findings, supervision of the study, and critically reviewed and edited the manuscript. RKC, AM, IK, and KSB: study design, data interpretation, drafting of the manuscript, and writing—review and editing. CS, MH, SR, MS, and SB: study design and writing—review and editing. All authors had full access to all the data in the study and had final responsibility for the decision to submit for publication. MU, TS and RKC accessed and verified the data.

## Acknowledgements

The study was funded by the Research Council of Norway through its Centres of Excellence scheme (project number 223269) and the University of Bergen (UiB), Norway, to the Centre for Intervention Science in Maternal and Child Health (CISMAC) and the Innlandet Hospital Trust, Lillehammer, Norway. We thank study participants and their families for their valuable time in the study; the staff of the Child Health Research Project who collected, supervised and entered data, including Prof Laxman Shrestha (principal investigator); and the staff of the Siddhi Memorial Hospital and Shyam Sunder Dhaubadel (founder of the hospital).

## Data sharing statement

Data is available on request. In order to meet ethical requirements for the use of confidential patient data, requests must be approved by the Nepal Health Research Council (NHRC) and the Regional Committee for Medical and Health Research Ethics in Norway. Requests for data should be sent to the authors, by contacting NHRC (http://nhrc.gov.np), or by contacting the Department of Global Health and Primary Care at the University of Bergen.

