## Supplementary table 1, Supplementary table 2, Supplementary figure 1 for "Women’s Body Mass Index trajectories from early pregnancy to one year postpartum and the rising burden of overweight and obesity over the last two decades in Bhaktapur, Nepal": Supplementary Appendix.pdf

**Supplementary Table 1.** Demographic characteristics of women from four studies in Bhaktapur, Nepal between 2001 and 2021.

| | **2001-2002** | **2007-2008** | **2015-2017** | **2017-2021** |
|---|---|---|---|---|
| **Total numbers of women** | 500 | 500 | 600 | 800 |
| Women's age (years), mean (SD) | 23 ± 6 | 25.8 ± 4.2 | 27.3 ±4.6 | 27.5± 4.0 |
| **Education**:<br>Completed school grade 10 or more above (≥10 grade) | 96 (20%) | 220 (47.4%) | 377 (62.8%) | 625 (78.1%) |
| **Occupation of women:**<br>Homemaker/Agriculture, n (%) | 205 (41%) | 363 (72.9%) | 373 (62.2%) | 283 (35.3%) |
| Family staying in joint family, n (%) | 258 (51.6%) | 251 (51%) | 292 (48.6%) | 522 (65.3%) |
| Mean number of family member | 6.5 (3.1) | | 5.3 (2.3) | 5 (2.5) |
| Mean number of rooms used by family | 3.1 (2.0) | | 1.6 (0.7) | 1.2 (0.4) |
| Family residing in rented house, n (%) | | | 291 (48.5%) | 198 (24.8%) |
| Kitchen and bedroom in the same room, n (%) | 159 (32.0%) | 182 (38%) | 298 (49.6%) | 216 (26.8%) |
| Family ownership of land, n (%) | 333 (66.6%) | | 282 (47.0%) | 539 (67.4%) |

Numbers are N (%) if not otherwise stated.

**Supplementary table 2.** Overweight and obesity among women from a longitudinal study in Bhaktapur, Nepal, 2017-2021, based on BMI by South Asian cut offs

| | BMI | | n (%) | |
|---|---|---|---|---|
| | | 1st trimester | 6 months PP | 12 months PP |
| Undernutrition | <18.5 | 0 | 4 (0.5%) | 7 (1.0%) |
| Normal | 18.5–22.9 | 348 (43.5%) | 127 (17.3%) | 185 (25.3%) |
| Overweight | 23–24.9 | 189 (23.6%) | 154 (21.0%) | 175 (23.9%) |
| Obesity | ≥25 | 263 (32.8%) | 449 (61.2%) | 364 (49.8%) |
| Grade I | 25–29.9 | 263 (32.8%) | 352 (48.0%) | 301 (41.2%) |
| Grade II | 30–34.9 | 0 | 95 (12.9%) | 60 (8.2%) |
| Grade III | ≥35 | 0 | 2 (0.3%) | 3 (0.4%) |

**Supplementary table 3.** Trends in BMI according to South Asian specific cut offs of women in childbearing age or during lactation in four different studies last 20 years in Bhaktapur, Nepal

| Year | Total Sample | Period | BMI Mean (SD) | BMI (%) | | | |
|---|---|---|---|---|---|---|---|
| | | | | <18.5 | 18.5-22.9 | 23-24.9 | ≥25 |
| 2018 - 2022 | 731 | Postpartum 6-12 months | 25.6 (±3.3) | 5 (0.7%) | 155 (21.3%) | 165 (22.5%) | 406 (55.5%) |
| 2015 – 2017 Third study | 600 | Postpartum 6-11 months | 23.7 (±3.6) | 34 (5.7%) | 223 (37.2%) | 134 (22.3%) | 209 (34.8%) |
| 2007 – 2008 Second study | 500 | Postpartum 2-11 months | 22.5 (±3.1) | 23 (4.6%) | 302 (60.2%) | 91(18.2%) | 85 (17%) |
| 2001 – 2002 First study | 500 | Childbearing age | 21.8 (±3) | 55 (11%) | 306 (61.2%) | 73 (14.6) | 66 (13.2%) |

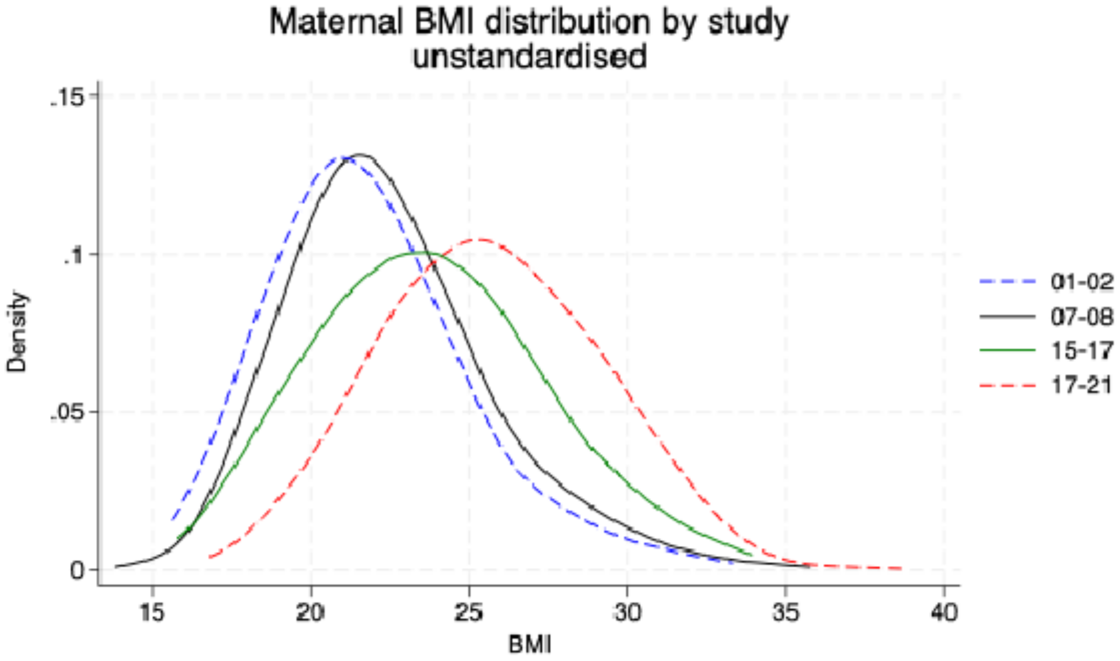


**Supplementary Figure 1.** Unadjusted distribution of women's BMI among participants from four studies conducted in Bhaktapur, Nepal (2000–2020).

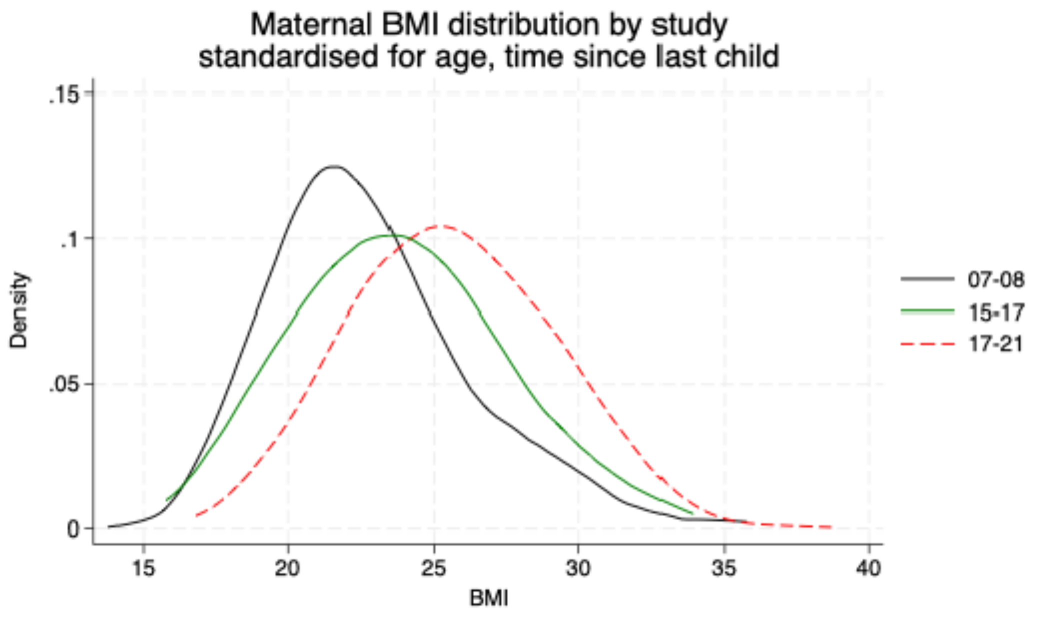


**Supplementary Figure 2.** Distribution of women's BMI adjusted for maternal age and child age across three studies conducted in Bhaktapur, Nepal (2000–2021)
